# Now-casting the COVID-19 epidemic: The use case of Japan, March 2020

**DOI:** 10.1101/2020.03.18.20037473

**Authors:** Stephan Glöckner, Gérard Krause, Michael Höhle

**Affiliations:** Helmholtz-Centre for Infection Research, Department of Epidemiology, Braunschweig, Germany; German Center for Infection Research (DZIF); Department of Mathematics, Stockholm University, Stockholm, Sweden

## Abstract

**Background:** Reporting delays in disease surveillance impair the ability to assess the current dynamic of an epidemic. In continuously updated epidemic curves, case numbers for the most recent epidemic week or day usually appear to be lower than the previous, suggesting a decline of the epidemic. In reality, the epidemic curve may still be on the rise, because reporting delay prevents the most recent cases to appear in the case count. In context of the COVID-19 epidemic and for countries planning large international gatherings, such as the Summer Olympic Games in Japan 2020, the ability to assess the actual stage of an epidemic is of outmost importance.

**Methods:** We applied now-casting onto COVID-19 data provided by the nCoV-2019 Data Working Group to evaluate the true count of cases, by taking into account reporting delays occurring between date of symptom onset and date of confirmation.

**Findings:** We calculated a decrease of reporting delay, from a median delay of ten days in calendar week four 2020 to six days in calendar week eight, resulting in an overall mean of 4.3 days. The confidence intervals of the now-casting indicated an increase of cases in the last reporting days, while case country in that same time period suggested a decline.

**Interpretation:** As a specific use case this tool may be of particular value for the challenging risk assessment and risk communication in the context of the Summer Olympic Games in Japan 2020 and similar situations elsewhere.

## Introduction

Reporting delay of cases is a common obstacle for real-time risk assessment and management of epidemics. In continuously updated epidemic curves, case numbers for the most recent epidemic week or day usually appear to be lower than the previous, suggesting a decline of the epidemic. In reality however, the epidemic curve may still be on the rise, because reporting delay prevents the most recent cases to already appear in the case count. This phenomenon often introduces misperceptions in the general public and in media coverage as can again be seen in the current COVID-19 epidemic.^1^

Reporting delay can be decomposed into several parts: (a) the time passing from symptom onset until the patient seeks medical service, (b) the time passing between this service and initiation of a laboratory test, (c) the time between this test initiation and completion of its result, (d) the time between test completion and its registration at the local health authority level and (e) the time passing from transmitting the case report from that local level to the (usually national) health authority level that will generate and publish the epidemic curve.^2^

Technology and rapid procedures can minimize the delay occurring from the first contact with the health care system and appearance of a case in the daily case count at national level, but they can hardly reduce the delay between appearance of first symptoms and contact to the health care service.^2^ The latter is subject to a variety of factors such as severity of the disease, public awareness for it, individual benefit to seek health services, accessibility to those services, some of which may also be subject to change during an epidemic. For all those reasons, reporting delay is not only specific to a country and its surveillance and health care system, but to the disease in questions and the phase of the epidemic.

For the COVID-19 in China, The World Health Organization (WHO) published a median reporting delay of 12 days (range 8-18 days) at the start of the outbreak and three days (range 1-7 days) in early February.^3^ Disease transmission models report a reporting delay larger than ten days during the start of the novel coronavirus outbreak and four days delay after the implementation of the quarantine procedures on January 27^th^.^4^ A study monitoring transmission potential of COVID-19 in Singapore estimated a reporting delay of 6.9 days.^5^ In contrast, the 2012 MERS-COV outbreak in Saudi-Arabia had a four day reporting delay^6^ and during the 2018 Measles outbreak, Japan showed a mean reporting delay of 4.5 days.^7^

Now-casting uses previous data to estimate the delay distribution while taking into account that cases with long-delays are currently not observed due to right-truncation with the aim to adjust epidemic curves for “observed-but-not-yet-reported cases”.^8,9^ On the occasion of the 2011 STEC/HUS outbreak in Germany the now-casting method was further optimized for the specific situation of being in the midst of an acute outbreak with interventions to actively reduce the delay.^10^

We report here on an improved concept of now-casting, that allows the adjustment of epidemic curves by taking into account that information about symptom onsets may not be available for a relevant proportion of the case reports. We demonstrate the approach with data from the COVID-19 outbreak in Japan, because of its high case counts, accessibility to data details and the particular importance of such an approach for the risk assessment for the upcoming Summer Olympic Games, hosted in Tokyo, July to August 2020. The aim of this approach is to reduce and transparently display uncertainty on the real time status of epidemic curves.

## Methods

For the purpose of this research, we define the reporting delay to be the time between the onset of disease symptoms of a case and the time the case report is available at the public health authority that generated the epidemic curve.^2^

We used the line-list dataset provided by the nCoV-2019 Data Working Group^11^ and WHO situation reports,^12^ to allow for a comparison between the two sources. On March 6^th^ 2020, nCoV-2019 Data Working Group contained 204 cases (excluding all cases from the Diamond Princess and with the last case reported on March 2^nd^ 2020), of which 131 (64%) contained date of symptom onset and date of case confirmation. For the remaining 73 (36%) cases without information on symptom onset, we assume that the case still had a symptom onset date and it is just missing, because it could not be readily identified. We imputed the respective date using a generalized additive Weibull regression model for both the scale and shape parameter with week of confirmation as a covariate.^13^

The now-casting approach consists of a Bayesian approach for modelling delay by which case reports appear in the continuously updated epidemic curve (also known as reporting triangle) and provides full predictive distributions for the daily delay-adjusted number of symptom onsets.^10^ We used a sliding window of the past 14 days to estimate the time-varying delay distribution, in order to account for the assumption that the reporting delay decreases as the outbreak progresses, due to aforementioned changes in the health care seeking and reporting procedures. Uncertainty of the delay imputation was transported by repeating the imputation 100 times and averaging the resulting now-casting quantities.

We developed this approach within R^14^ using the open-source R package surveillance^15^ and developed a Shiny application^16,17^, allowing users to perform now-casting without need for sophisticated programming skills and made it available at: https://helmholtz-epid.shinyapps.io/nowcasting_dashboard/.

## Results

The comparison of the nCoV-2019 Data Working Group line-list data and the data gathered from the WHO Situation reports for date of confirmation shows similar trends, while the WHO data seems to lag one day behind, starting February 14, 2020. WHO reported for February 5^th^ a cumulative total of 33 cases, but set the case count back on February 6^th^ to a cumulate total of 25 cases. We set the cases of February 6^th^ to unknown and restarted the plotting of the epidemic curve at this day (see Figure *1*).

Our modelling of symptom onsets show a decrease of reporting delay over the time (see Figure 2), from a median of ten days in calendar week four to six days in calendar week eight.

**Figure 1.**
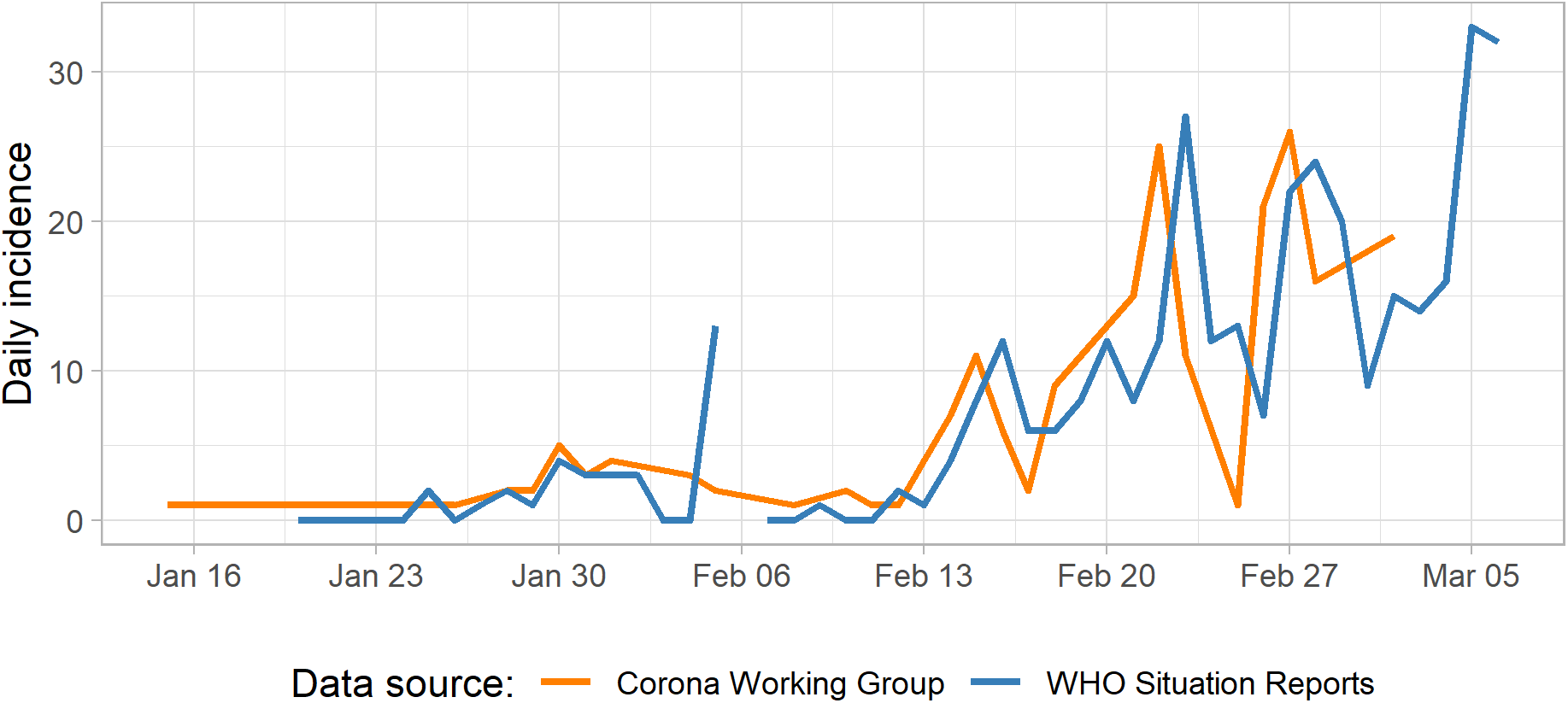
COVID-2019 Epidemic curve of confirmed cases for Japan as of March 6^th^ 2020, comparing data from nCoV-2019 Data Working Group and WHO.

**Figure 2.**
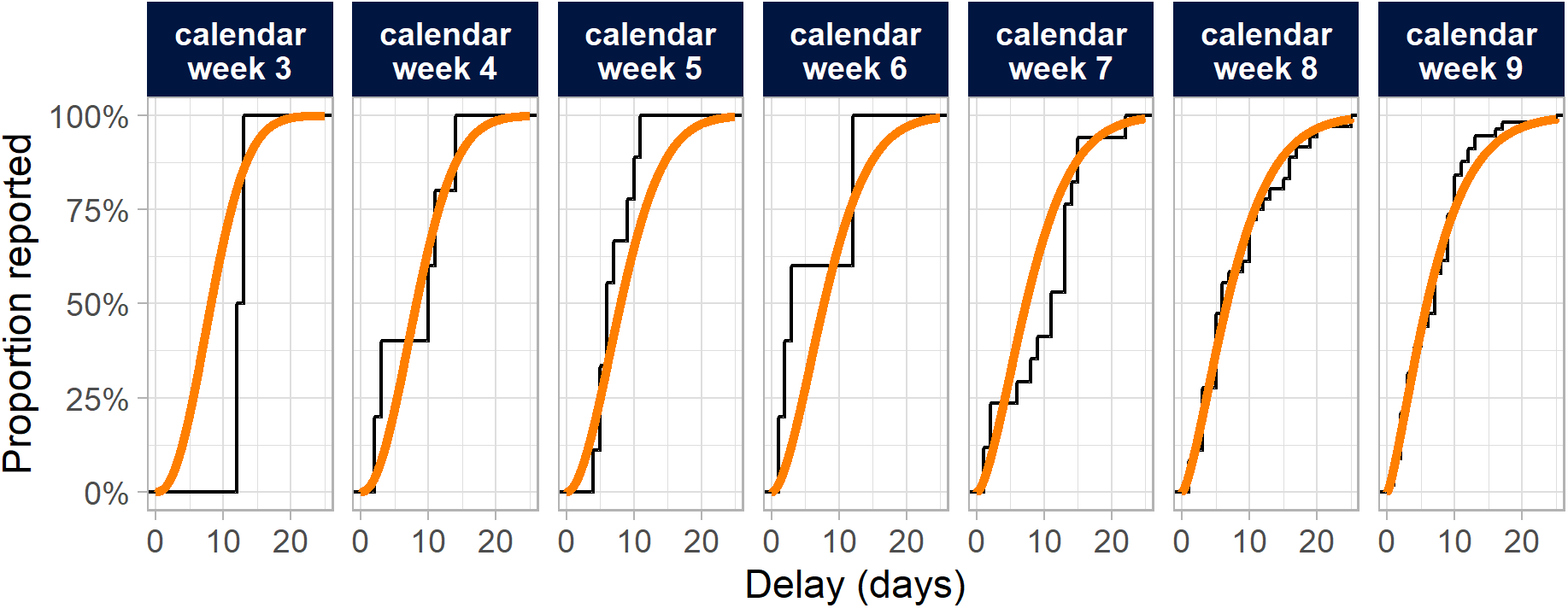
Empirical (black line) and model fitted (orange line) cumulative distribution function for the reporting delay for each calendar week.

Figure *3* shows the results of the now-cast for the line-list data on March 2^nd^ 2020 and done for the previous 14 days. Case counts more than nine days ago receive little adjustment, because the delay distribution is such that almost all cases are available. As an example, the number of now-casted cases on February 29^th^ 2020 is 6.2 on top of the observed 1.9 with a 95% prediction interval of 0.5 to 23.2. From the adjusted cases between February 28^th^ and March 1^st^ it becomes clear that despite the observed counts being low there appears to be an increasing trend. This trend is confirmed by the WHO data on confirmed cases for March 2^nd^ and beyond.

## Discussion

Our approach helps to assess whether the apparent decline is imminent to any real-time display of epidemic curves, is likely to reflect a true decline or more likely to reflect an artefact resulting from reporting delays. This artifact is immanent to any surveillance system and its resulting epidemic curves if continuously updating all data available at the time of publication. This approach also visualizes easily the level of uncertainty with this regard and therefore facilitates transparent public health risk communication.

In contrast to prediction models aiming to estimate the future development of an outbreak, the now-cast approach has the more humble, but no less valuable, objective to estimate whether an outbreak is still on the rise or already declining. In comparison to previous now-casting approaches, the method presented here, takes into account that a) reporting delay changes over time during an epidemic and also that b) for a relevant proportion of case reports not all data are available for computation of case specific reporting delays.

Public awareness is likely to have a major impact on surveillance sensitivity and also timeliness^18^: The WHO reported the first Corona case in Japan on January 20^th^ 2020, with four additional cases on January 25^th^. During this time, the media had already reported intensively about COVID-19 and therefore public awareness was likely to have been already high. When the first case of SARS-CoV-2 was reported in China on December 1^st^ 2019, the Japanese National Institute of Diseases established a real time PCR for this type of virus until January 16^th^ 2020. Furthermore, a telephone hotline for professionals within the health system and public communications were activated.^19^ These and other activities may explain why the reporting delay in Japan was fairly short to begin with and diminished further as the outbreaks progressed, compared to COVID-19 outbreaks in other countries.^3^ The delays displayed for calendar week nine as part of the imputation might experience a small downwards bias due to cases with long delays not being available at the date of query, however, this is then later taken into account by now-casting.

The added value of this now-casting approach increases with the length of the reporting delay – the larger the delays the more important is the adjustment. As illustrated per 95% prediction intervals in figure 3, uncertainty of now-casts is obviously highest for the days are closest to the actual day of the now-cast itself. We used publicly available data for our analysis, which tends to be less complete than the data available to national surveillance agencies. This slightly limits the precision of our now-cast, but also indicates the even higher value of now-casting when used within a nation’s surveillance system. For this reason, we are currently building this tool as a permanent functionality into the dashboard of the Surveillance Outbreak Response Management System (www.sormas.org) to further enhance its role in COVID-19 response.^20–23^

**Figure 3.**
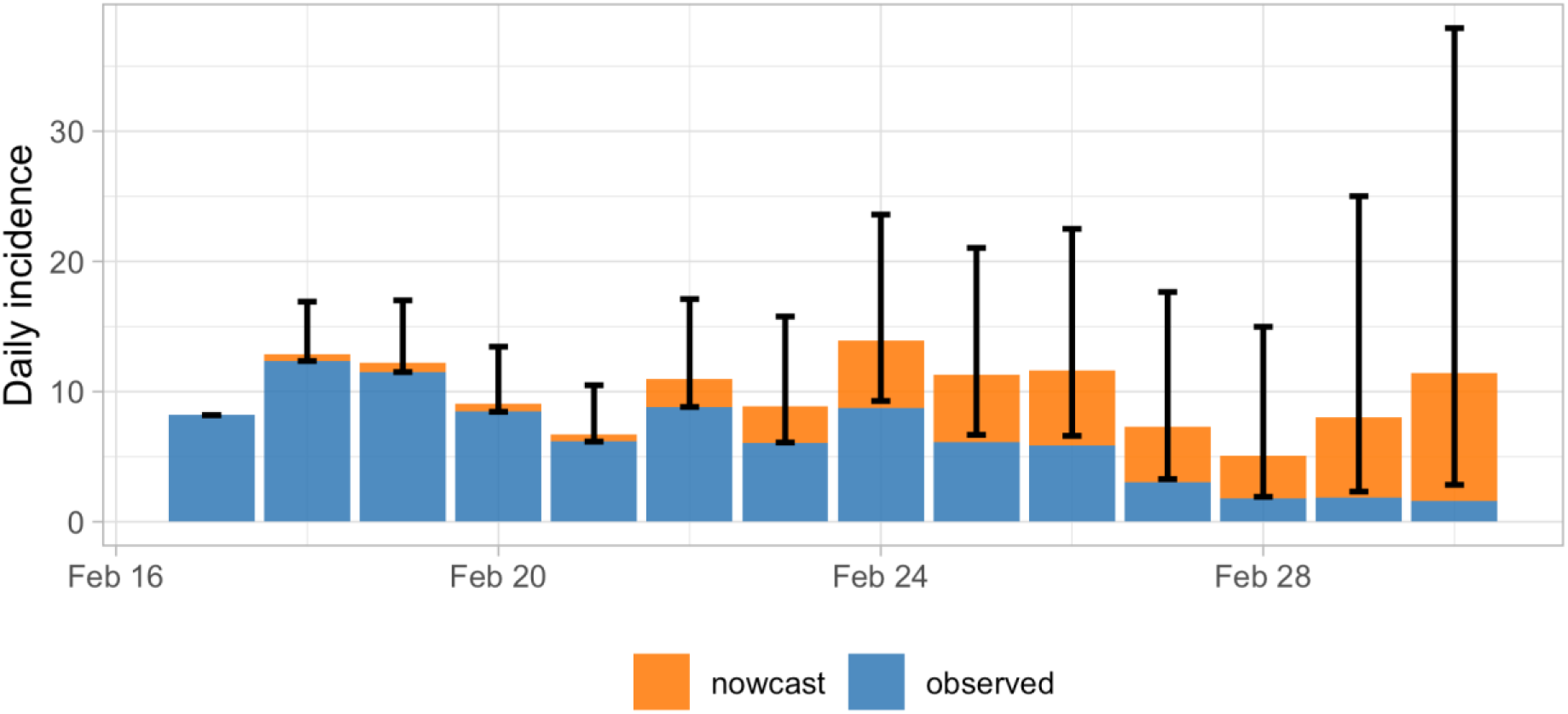
Now-cast for the epidemic curve of COVID-19 symptom onsets in Japan performed on March 2^nd^ 2020 for the previous 14 days. The orange area indicates the median of the predictive distribution for the number of cases missing. Furthermore, 95% prediction intervals for the total counts are shown.

Via the open-source R package surveillance applied within the user-friendly shiny application, public health surveillance institutions can easily apply this now-cast approach in their country for COVID-19 or any other disease. No system or country specific configurations are needed. As a specific use case this tool may be of particular value for the challenging risk assessment and risk communication in the context of the Summer Olympic Games in Tokyo July to August 2020 and similar situations elsewhere.

## Data Availability

We (the authors) had access to all data at all stages of our research. We will include links and data that will make our research reproducable for others.

https://github.com/gstephan30/nowcasting_COVID19

https://helmholtz-epid.shinyapps.io/nowcasting_dashboard/

## Acknowledgements

We thank the Japanese health authorities for generating and publishing surveillance data in the detail and timeliness necessary for the analyses in this report. In that regard, we thank the nCoV-2019 Data Working Group who gathered the Japanese case data from various online resources. The work of MH was supported by the Swedish Research Council (2015_05182_VR). Code be found on https://github.com/gstephan30/nowcasting_COVID19.

